# Characterizing the Clinical and Genetic Spectrum of Polycystic Ovary Syndrome in Electronic Health Records

**DOI:** 10.1101/2020.05.08.20095786

**Authors:** Ky’Era V. Actkins, Kritika Singh, Donald Hucks, Digna R. Velez Edwards, Melinda Aldrich, Jeeyeon Cha, Melissa Wellons, Lea K. Davis

**Affiliations:** Department of Microbiology, Immunology, and Physiology, Meharry Medical College, Nashville, TN; Vanderbilt Genetics Institute, Vanderbilt University Medical Center, Nashville, TN; Division of Genetic Medicine, Department of Medicine, Vanderbilt University Medical Center, Nashville, TN; Department of Thoracic Surgery, Vanderbilt University Medical Center, Nashville, TN; Department of Biomedical Informatics, Vanderbilt University Medical Center, Nashville, TN; Vanderbilt Epidemiology Center, Institute of Medicine and Public Health, Vanderbilt University Medical Center, Nashville, TN; Division of Quantitative Sciences, Department of Obstetrics and Gynecology, Vanderbilt University Medical Center, Nashville, TN; Division of Diabetes, Endocrinology, and Metabolism, Department of Medicine, Vanderbilt University Medical Center, Nashville, TN

**Keywords:** polycystic ovary syndrome, phenotyping, hormones, electronic health record, polygenic risk scores

## Abstract

**Context:** Polycystic ovary syndrome (PCOS) is one of the leading causes of infertility, yet current diagnostic criteria are ineffective at identifying patients whose symptoms reside outside strict diagnostic criteria. As a result, PCOS is under diagnosed and its etiology is poorly understood.

**Objective:** We aim to characterize the phenotypic spectrum of PCOS clinical features within and across racial and ethnic groups.

**Methods:** We developed a strictly defined PCOS algorithm (PCOS_regex-strict_) using International Classification of Diseases, 9^th^ and 10^th^ edition (ICD9/10) and regular expressions mined from clinical notes in electronic health records (EHRs) data. We then systematically relaxed the inclusion criteria to evaluate the change in epidemiological and genetic associations resulting in three subsequent algorithms (PCOS_coded-broad_, PCOS_coded-strict_,PCOS_regex-broad_). We evaluated the performance of each phenotyping approach and characterized prominent clinical features observed in racially and ethnically diverse PCOS patients.

**Results:** The best performing algorithm was our PCOS_coded-strict_ algorithm with a positive predictive value (PPV) of 98%. Individuals classified as cases by this algorithm had significantly higher body mass index (BMI), insulin levels, free testosterone values, and genetic risk scores for PCOS, compared to controls. Median BMI was higher in African American women with PCOS compared to White and Hispanic women with PCOS.

**Conclusions:** PCOS symptoms are observed across a severity spectrum that parallels genetic burden. Racial and ethnic group differences exist in PCOS symptomology and metabolic health across different phenotyping strategies.

## Introduction

Polycystic ovary syndrome (PCOS) is an endocrine disorder that is the leading cause of infertility in women. The genetic, environmental, and metabolic variables that contribute to its complex architecture also influence clinical heterogeneity among individuals with PCOS, resulting in a broad spectrum of symptoms. The heterogenous clinical presentation of PCOS makes it difficult to diagnose and an estimated 75% of women with PCOS remain undiagnosed (1,2).

There are three commonly used diagnostic criteria for PCOS which each have specific symptom requirements for diagnosis. The first diagnostic definition was put forth by the National Institute of Health (NIH) who required an ovulatory phenotype (i.e., oligomenorrhoea) and hyperandrogenism for diagnosis (3,4). Subsequently, the second diagnostic definition for PCOS was created by the European Society for Human Reproduction and Embryology and the American Society for Reproductive Medicine, which is commonly referred to as Rotterdam criteria. The Rotterdam diagnostic criteria was designed to expand the PCOS definition from the strict NIH definition by requiring any two out of the following three symptoms: oligoanovulation, hyperandrogenism, or polycystic ovaries (3,5). However, the Androgen Excess and PCOS Society argued that hyperandrogenism is the primary driver of PCOS and thus, created a third diagnostic criteria that requires both hyperandrogenism and ovulatory dysfunction (i.e., oligoanovulation or polycystic ovaries) (4). PCOS diagnostic criteria were developed to improve and standardize diagnosis, however, patients identified by each approach may have differing symptomologies and only a small fraction of women will meet multiple diagnostic criteria (6,7).

Implementing a broader PCOS definition could lead to false positive diagnoses and prevalence overestimations (4,8). However, restrictive definitions miss women with clinical manifestations outside of the requirements (9) including metabolic dysfunction. Women with PCOS and metabolic dysfunction are at higher risk for a range of comorbidities including insulin resistance, type 2 diabetes, cardiovascular disease, obesity, cancer, and psychiatric disorders (10,11). Delayed screening <u>and</u> treatment for PCOS could exacerbate these metabolic conditions, particularly in underrepresented populations who have a greater baseline risk of metabolic disorders (12,13). Studies have shown that African American and Hispanic women with PCOS have an increased risk for metabolic syndrome and other cardiovascular disorders, yet little is known about why comorbidities increase across diverse racial and ethnic groups and whether expanding diagnostic criteria for PCOS may improve health equity (14,15).

Most prior studies included small numbers of minority women and many have used differing diagnostic criteria (1,16–18). Without larger studies, it is difficult to accurately characterize the clinical presentation of PCOS and its consequences across populations. Adapting an inclusive phenotyping approach capable of identifying severe to mild PCOS presentations could improve study methods for marginalized communities.

The overarching hypothesis of this study is that clinically diagnosable PCOS represents the extreme tail (i.e., clinical manifestation) of a spectrum of hormonal and metabolic dysregulation with common manifestations in a large number of women. In our study, we developed a simple but stringent automated phenotyping algorithm that could be applied to electronic health records (EHRs) to identify women with confirmed PCOS. Next, we systematically relaxed the inclusion criteria to test the hypothesis that excess androgens, increased metabolic dysfunction, and PCOS genetic liability are present to a lesser degree among women with PCOS symptoms outside diagnostic criteria. Finally, we examined these features across diverse racial and ethnic groups within the EHR.

## Methods

### Clinical data

Vanderbilt University Medical Center (VUMC) is a tertiary care hospital in Nashville, Tennessee, with several clinics throughout Tennessee and the surrounding states. Medical records have been electronically documented at VUMC since the early 1990s (19). The VUMC EHR houses over 2.8 million EHRs in a de-identified clinical research database referred to as the Synthetic Derivative (SD). The SD includes demographics, International Classification of Diseases, Ninth Revision (ICD9) and Tenth Revision (ICD10) codes, procedural codes (CPT), clinical notes, medications, and laboratory values. The Vanderbilt University Institutional Review Board approved this project (IRB 160279).

### Algorithm development

We identified women with PCOS within the SD using a strict automated phenotyping algorithm that yielded a positive predictive value (PPV) of at least 90%.

Our secondary analyses relaxed the requirements of our core algorithm to identify a larger sample of patients with a wider array of PCOS symptoms. In total, we developed four algorithms which are illustrated in **Figure 1**. The demographics of each resultant dataset is reported in **Table 1**.

**Figure 1.**
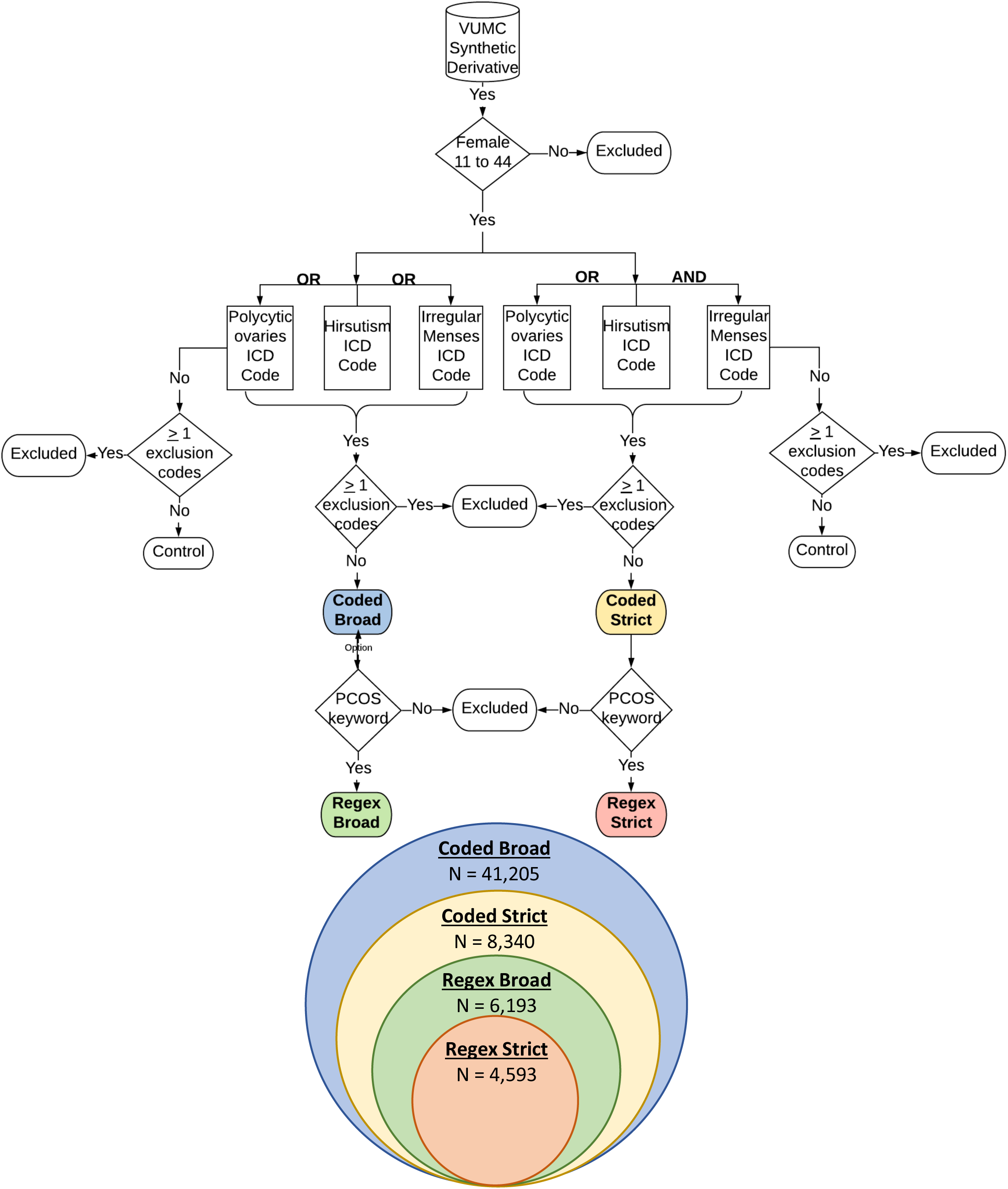
PCOS algorithm pipeline. This schematic illustrates the algorithm development steps for the controls and the four PCOS algorithms. The Venn diagram illustrates how the algorithm datasets are nested within one another. Each color corresponds with one of the algorithm datasets. ICD = International classification of disease, 9^th^ and 10^th^ revision.

**Table 1.** PCOS algorithm descriptive statistics. Descriptive statistics of women identified as PCOS cases and controls are provided. SD = standard deviation; EHR = electronic health record. NH = non-Hispanic, HIS = Hispanic.

|  | Controls | Coded Broad | Coded Strict | Regex Broad | Regex Strict |
| --- | --- | --- | --- | --- | --- |
| N | 29140 | 41205 | 8340 | 6193 | 4593 |
| NH White | 22569 | 25977 | 5418 | 4154 | 3101 |
| NH African American | 3278 | 7555 | 1195 | 887 | 648 |
| Hispanic | 1421 | 2053 | 368 | 274 | 204 |
| HIS White | 1014 | 1462 | 264 | 203 | 154 |
| HIS African American | 33 | 69 | 13 | 11 | 8 |
| Average Age (SD) | 23.85<br>(9.47) | 24.96<br>(7.97) | 25.18<br>(7.63) | 24.71<br>(7.52) | 25.09<br>(7.39) |
| Average EHR Record Length (SD) | 3.92<br>(3.9) | 4.52<br>(4.54) | 4.23<br>(4.15) | 4.69<br>(4.22) | 4.66<br>(4.13) |
| Medical Home (%) | 65% | 64% | 60% | 66% | 66% |
| Average Clinical Values (SD) |  |  |  |  |  |
| Body Mass Index | 25.71<br>(7.43) | 28.61<br>(8.44) | 34.12<br>(9.10) | 33.50<br>(9.14) | 34.54<br>(9.07) |
| Insulin (mcU/mL) | 20.08<br>(18.99) | 24.86<br>(22.83) | 27.17<br>(23.05) | 25.34<br>(21.94) | 27.27<br>(21.58) |
| Estradiol (pg/mL) | 82.22<br>(69.03) | 89.92<br>(69.74) | 75.92<br>(61.13) | 71.64<br>(56.56) | 75.57<br>(60.49) |
| Free Testosterone (pg/mL) | 3.58<br>(2.45) | 4.70<br>(2.94) | 5.70<br>(3.23) | 5.41<br>(3.01) | 5.73<br>(3.09) |

Before applying each algorithm, we filtered the entire SD population to include only females between the ages of 11 to 44 to enrich our population for females of reproductive age. We first deployed the PCOS_regex-strict_ algorithm which required at least one ICD9 or ICD10 code for: polycystic ovaries (i.e., 256.4 and E28.2) OR hirsutism (i.e., 704.1 and L68.0) AND irregular menses (i.e., all 626 codes, all N91 codes, N92.5, and N92.6) **(Supplementary Table 1)**. Hirsutism was used as clinical evidence of hyperandrogenism. Individuals with any exclusion codes **(Supplementary Table 1)** present in their medical record were removed. Furthermore, the PCOS_regex-strict_ algorithm required the presence of at least one of the following regular expressions, “polycystic ovaries”, “PCOS”, or “PCO” in a clinical note. We included inpatient and outpatient note types designated as “Outpatient note”, “Gynecology clinic visit”, “Nursing report”, “Endocrine and Diabetes Clinic Visit”, “Pediatric Endocrinology Patient Visit”, and “Reproductive Endocrinology Clinic Visit”. Next, to broaden the approach, we dropped the regular expression requirements and retained only the ICD codes for the PCOS_coded-strict_ algorithm. We then loosened the requirement of the union of ICD codes and instead required the presence of ICD codes for polycystic ovaries <u>OR</u> hirsutism <u>OR</u> irregular menses. In addition to any code for these conditions, we required the presence of a PCOS-related regular expression. The resultant algorithm was termed “PCOS_regex-broad_”. Lastly, we again dropped the regular expression requirement which resulted in the broadest approach (PCOS_coded-broad_) including only the presence of ICD codes for polycystic ovaries <u>OR</u> hirsutism <u>OR</u> irregular menses.

To identify controls, the SD was again filtered for females between the ages of 11 to 44. The control dataset excluded individuals with one or more exclusion ICD codes or any inclusion codes used to define PCOS case criteria **(Supplementary Table 1)**.

### Chart review methods for algorithm evaluation

We performed two comprehensive chart reviews to evaluate the performance of the phenotyping algorithms. The charts were reviewed by clinical domain experts (MW and JC) and trained reviewers (KVA, DH, and KS). We first evaluated the positive predictive value (PPV) for each algorithm. We randomly selected 50 charts from each set of algorithm defined cases (200 in total) for manual chart review. We then calculated the proportion of true positive cases based on manual chart review out of the total algorithm identified cases, to define the PPV for each algorithm (**Table 2**).

**Table 2.** Prevalence for PCOS algorithms and chart review results. Prevalence of PCOS was calculated for everyone identified in each algorithm (overall) and for White, African American, and Hispanic women identified in each algorithm. Chart review was conducted on 50 random records identified by each PCOS algorithm (n = 200) and algorithm performance was measured through PPV. PPV = positive predictive value.

|  | Coded<br>Broad | Coded<br>Strict | Regex<br>Broad | Regex<br>Strict |
| --- | --- | --- | --- | --- |
| Overall |  |  |  |  |
| N | 41205 | 8340 | 6193 | 4593 |
| Prevalence | 46% | 9.3% | 6.9% | 5.1% |
| PPV | 30% | 98% | 82% | 96% |
| White |  |  |  |  |
| N | 26317 | 5526 | 4207 | 3143 |
| Prevalence | 50% | 10.5% | 8% | 4.8% |
| African American |  |  |  |  |
| N | 26317 | 5526 | 4207 | 3143 |
| Prevalence | 50% | 10.5% | 8% | 4.8% |
| Hispanic |  |  |  |  |
| N | 2052 | 368 | 274 | 204 |
| Prevalence | 48% | 8.6% | 6.4% | 4.8% |

Next, we evaluated the sensitivity and specificity of the original PCOS_regex-strict_ algorithm. For this review, we first identified a “medical home” population (defined by the presence of at least 5 ICD codes on separate days over at least a 3-year period (N = 752,436)). Then, we filtered for females who met a data floor requirement (i.e., at least one ICD10 code for PCOS, E28.2 (N = 9,507)). We then selected 200 charts at random and performed manual chart review of all available information in the EHR (i.e., codes and clinical notes) to classify individuals into “PCOS cases” and “not PCOS cases”. This manual review clinical determination was then compared to the PCOS_regex-strict_ algorithm-defined determination to calculate sensitivity and specificity (**Table 3**).

**Table 3.** Sensitivity and specificity for PCOS algorithm datasets. A 2×2 contingency table showing the PPV, NPV, sensitivity, and specificity of the regex strict (RS) algorithm. The RS algorithm was compared to the gold standard definition. Chart review was conducted on 200 random records from females who met our data floor and medical home definition. PPV = positive predictive value; NPV = negative predictive value.

|  |  | Regex Strict Algorithm |  |  |
| --- | --- | --- | --- | --- |
|  |  | Case | Not Case | Results |
| Gold Standard | Case | 96 | 97 | 50% Sensitivity |
|  | Not case | 5 | 2 | 29% Specificity |
|  | Results | 95% PPV | 2% NPV |  |

### Laboratory data

Weight and height measurements are routinely recorded during clinical visits and were extracted from the EHR to calculate body mass index (BMI). Laboratory measurements were extracted and cleaned using QualityLab (20). In brief, 3,075 quantitative laboratory tests were extracted from EHRs of 70,704 patients in the SD. Laboratory values with non-numeric values (n=2,865) and labs evaluated on only one patient (n=467) were removed. The remaining labs were cleaned by filtering out duplicate entries and measurements with multiple units. Insulin, estradiol, and free testosterone hormone data was then extracted for further analysis. Median laboratory values were calculated for individuals with multiple clinical measurements. Median laboratory values of BMI, insulin, estradiol, and free testosterone were used to characterize the clinical features of each algorithm dataset (**Table 4 and Supplementary Table 2**). For each lab, extreme outliers (i.e., 1^st^ and 99^th^ percentile) were removed. Additionally, lab values were regressed on age at lab and an inverse normal transformation was applied to the residual to result in an age-adjusted normalized lab value for subsequent analysis including genetic analysis (**Supplementary Table 5**). Sample sizes differed for subsequent analyses based on available clinical measurements (**Supplementary Figure 1**).

**Table 4.** Laboratory measurements for PCOS algorithm cases and controls. Wilcoxon rank sum test was performed between cases and controls. The 25^th^ and 75^th^ percentile was reported for each algorithm. BMI = body mass index.

| Algorithm | Type | N | Median | 25th Percentile | 75th Percentile | P |  |
| --- | --- | --- | --- | --- | --- | --- | --- |
| BMI |  |  |  |  |  |  |  |
| Coded Broad | Cases | 34603 | 26.54 | 22.21 | 33.50 | < 2.22E-16 | * * |
|  | Controls | 29140 | 24.03 | 20.68 | 29.30 |  |  |
| Coded Strict | Cases | 7555 | 33.605 | 27.28 | 40.36 | < 2.22E-16 | * * |
|  | Controls | 29140 | 24.03 | 20.68 | 29.30 |  |  |
| Regex Broad | Cases | 5844 | 32.805 | 26.29 | 39.70 | < 2.22E-16 | * * |
|  | Controls | 29140 | 24.03 | 20.68 | 29.30 |  |  |
| Regex Strict | Cases | 4358 | 34.08 | 27.63 | 40.76 | < 2.22E-16 | * * |
|  | Controls | 29140 | 24.03 | 20.68 | 29.30 |  |  |
| Insulin |  |  |  |  |  |  |  |
| Coded Broad | Cases | 257 | 17.4 | 10.00 | 24.10 | 0.0001 | * * |
|  | Controls | 14475 | 13.65 | 5.50 | 20.90 |  |  |
| Coded Strict | Cases | 150 | 19.05 | 11.20 | 28.70 | 1.50E-06 | * * |
|  | Controls | 14475 | 13.65 | 5.50 | 20.90 |  |  |
| Regex Broad | Cases | 134 | 18.1 | 10.52 | 26.06 | 0.00088 | * |
|  | Controls | 14475 | 13.65 | 5.50 | 20.90 |  |  |
| Regex Strict | Cases | 109 | 19.4 | 11.53 | 32.53 | 2.40E-05 | * * |
|  | Controls | 14475 | 13.65 | 5.50 | 20.90 |  |  |
| Estradiol |  |  |  |  |  |  |  |
| Coded Broad | Cases | 196 | 63.5 | 38.00 | 116.00 | 0.0081 |  |
|  | Controls | 18799 | 55.5 | 32.00 | 108.00 |  |  |
| Coded Strict | Cases | 67 | 54 | 38.00 | 96.65 | 0.79 |  |
|  | Controls | 18799 | 55.5 | 32.00 | 108.00 |  |  |
| Regex Broad | Cases | 58 | 54.5 | 38.50 | 90.88 | 0.86 |  |
|  | Controls | 18799 | 55.5 | 32.00 | 108.00 |  |  |
| Regex Strict | Cases | 48 | 55.5 | 39.50 | 93.20 | 0.64 |  |
|  | Controls | 18799 | 55.5 | 32.00 | 108.00 |  |  |
| Free Testosterone |  |  |  |  |  |  |  |
| Coded Broad | Cases | 159 | 4.4 | 2.25 | 6.58 | 0.0016 | * |
|  | Controls | 125 | 3 | 1.70 | 4.95 |  |  |
| Coded Strict | Cases | 75 | 5.9 | 3.00 | 7.70 | 3.80E-06 | * * |
|  | Controls | 125 | 3 | 1.70 | 4.95 |  |  |
| Regex Broad | Cases | 73 | 5.2 | 2.90 | 7.40 | 3.50E-05 | * * |
|  | Controls | 125 | 3 | 1.70 | 4.95 |  |  |
| Regex Strict | Cases | 53 | 5.6 | 3.60 | 7.50 | 1.40E-05 | * * |
|  | Controls | 125 | 3 | 1.70 | 4.95 |  |  |

### Genetic data

We included 40,006 females of European descent and 7,736 females of non-European descent who were genotyped on the Illumina MEGA^EX^ Array (Illumina Inc, San Diego, CA) (21). We removed SNPs with a call rate < 0.98, individuals with a call rate of < 0.98, and individuals with a discrepancy between chromosomal and reported sex. EIGENSTRAT was used for principal component analysis (PCA) (22,23). Related samples with a pi-hat > 0.2 (approximate first cousin relationships) were removed. The Michigan Imputation server was used to impute missing genotypes based on the Haplotype Reference Consortium panel and imputation accuracy was set at R^2^>0.03 (24).

### Construction of PCOS polygenic risk scores (PCOS_PRS_)

Polygenic risk scores (PRS) for PCOS were calculated for each individual using the PRS-CS software package, which applies a Bayesian continuous shrinkage model to SNP beta estimates (25). Beta estimates were drawn from summary statistics of the largest meta-GWAS of PCOS performed by Day et al. which included 5,209 cases and 32,055 controls of European descent diagnosed using NIH or Rotterdam criteria (26). *PCOS_PRS_* were calculated for a total of 36,487 females of European descent, 5,582 non-European descent females, and 29,511 females from multiple ancestries in BioVU.

### Statistical Analysis

The prevalence of each algorithm was calculated using the proportion of cases identified by each algorithm among total number of EHRs (**Table 2**). The PPV was determined by calculating the proportion of “reviewer-defined” true positives among “algorithm-defined” positives (**Table 2**). Sensitivity was defined as the number of “algorithm-defined” positives out of the total positives (i.e., “algorithm-defined” positives plus “reviewer-defined” positives). Specificity was defined as the number of “algorithm-defined” negatives out of the total negatives (i.e., “algorithm defined and “reviewer-defined” negatives) (**Table 3**).

We performed Wilcoxon rank-sum tests to determine if the median BMI and laboratory measurements (e.g., free testosterone, insulin, and estradiol) differed between the PCOS algorithm-identified cases and controls. We then tested the difference between cases defined by each algorithm and between cases and controls stratified by EHR-reported race. Finally, we evaluated differences between African American, White, and Hispanic women identified as cases by each algorithm (**Table 4, Supplementary Table 2, Figures 2-4, Supplementary Figures 2-3**).

We performed logistic and linear regression analyses to test the association between PCOS_PRS_ and case/control designation and, separately, the association between PCOS_PRS_ and available hormone data (i.e., estradiol, free testosterone, insulin). The analysis was performed in three populations based on genetic ancestry: European descent (n=6,930), non-European (n = 2,050), and multi-ancestry (n=7,135). Our multi-ancestry cohort includes all genotyped individuals (European and non-European descent). Covariates included median age (calculated across the medical record for each individual) and top ten principal components of ancestry for the case/control logistic regression model. A separate sensitivity analysis adjusting BMI was also performed (**Supplementary Table 3**). For regression analysis of hormone data, covariates included the top ten principal components of ancestry. We then evaluated the model and variance explained by PCOS_PRS_ using Nagelkerke’s pseudo R^2^ for the logistic regressions and R^2^ for the linear regressions (**Table 5**, **Supplementary Table 3-4**). All of the analyses were done in R 3.3.3.

**Table 5.** Genetic validation of PCOS diagnosis. Logistic regressions were performed between PCOS case status and PCOS polygenic risk scores (PCOS_PRS_). The regressions were adjusted for median age and the first 10 principal components. SE = standard error; OR = odds ratio; LCI = lower confidence interval; UCI = upper confidence interval.

| Algorithm | Cases | Controls | Estimate | SE | P | OR | LCI | UCI | r<sup>2</sup> |  |
| --- | --- | --- | --- | --- | --- | --- | --- | --- | --- | --- |
| European |  |  |  |  |  |  |  |  |  |  |
| Coded Broad | 1878 | 5052 | 0.04 | 0.03 | 0.235 | 1.04 | 0.98 | 1.10 | 0.0003 |  |
| Coded Strict | 366 | 5052 | 0.19 | 0.06 | 0.002 | 1.21 | 1.07 | 1.36 | 0.0046 | * |
| Regex Broad | 332 | 5052 | 0.26 | 0.06 | 4.6E-5 | 1.29 | 1.14 | 1.46 | 0.0083 | ** |
| Regex Strict | 245 | 5052 | 0.21 | 0.07 | 0.004 | 1.23 | 1.07 | 1.42 | 0.0049 |  |
| Non-European |  |  |  |  |  |  |  |  |  |  |
| Coded Broad | 741 | 1309 | 0.09 | 0.10 | 0.354 | 1.10 | 0.90 | 1.33 | 0.0006 |  |
| Coded Strict | 126 | 1309 | 0.32 | 0.20 | 0.111 | 1.38 | 0.93 | 2.04 | 0.0039 |  |
| Regex Broad | 103 | 1309 | 0.28 | 0.22 | 0.208 | 1.32 | 0.86 | 2.04 | 0.0027 |  |
| Regex Strict | 77 | 1309 | 0.21 | 0.25 | 0.404 | 1.24 | 0.75 | 2.03 | 0.0014 |  |
| Multi-Ancestry |  |  |  |  |  |  |  |  |  |  |
| Coded Broad | 2228 | 4907 | 0.08 | 0.05 | 0.114 | 1.08 | 0.98 | 1.20 | 0.0005 |  |
| Coded Strict | 419 | 4907 | 0.40 | 0.10 | 8.7E-5 | 1.49 | 1.22 | 1.81 | 0.0068 | ** |
| Regex Broad | 382 | 4907 | 0.34 | 0.10 | 0.001 | 1.41 | 1.15 | 1.73 | 0.0050 | * |
| Regex Strict | 281 | 4907 | 0.36 | 0.12 | 0.003 | 1.44 | 1.13 | 1.82 | 0.0050 |  |

We included two corrections for multiple testing. The first corrected threshold reflects adjustment for each of the independent tests (n = 30, p < 0.002, indicated by * in tables). The second corrected threshold is more stringent and reflects adjustment for each of the tests (n = 120, p < 4×10^-3^, indicated by ** in tables) regardless of whether they were independent or performed on nested subsets of the data (**Figure 1**).

## Results

### Sample Characteristics

PCOS dataset demographics are reported in **Table 1**. Although the sample sizes differed between the algorithms, each algorithm identified a similar proportion of individuals across racial/ethnic groups. The most stringent algorithm, PCOS_regex-strict_, identified 4,593 potential PCOS cases including 3,143 White, 658 African American and 204 Hispanic women (**Table 2**). As the algorithm requirements were relaxed, the number of identified PCOS cases increased. PCOS_regex-broad_ identified 6,193 individuals including 4,207 White, 899 African American, and 274 Hispanic women. The PCOS_coded-strict_ approach identified 8,340 individuals including 5,526 White, 1,217 African American, and 368 Hispanic women. At the broadest extreme, the PCOS_coded-broad_ (n = 41,205) algorithm captured the largest and most diverse sample with 26,317 White, 7,648 African American, and 2,052 Hispanic women.

The individuals identified by the regex algorithms had the highest average medical record length at 4.69 years (SD = 4.22) for PCOS_regex-broad_ and 4.66 years (SD = 4.13) for PCOS_regex-strict_. This was expected as the increased length of record also increased the probability of PCOS documentation in the medical record. The PCOS_coded-strict_ and PCOS_regex-strict_ algorithms captured the eldest set of patients who were in their mid 20s with a median age of 25.18 (SD = 7.63) and 25.09 (SD = 7.39), respectively. Comparatively, the control dataset had the youngest sample aged 23.85 (SD = 9.47) with the shortest EHR record length at 3.92 (SD = 3.9). As expected, the control sample had the lowest average clinical measurements for BMI, insulin, estradiol, and free testosterone.

### Evaluation of algorithm performance

We found little difference between the PPVs calculated for the PCOS_regex-strict_ algorithm using two distinct chart review methods. PCOS_regex-strict_ yielded a PPV of 96% using our first method of chart review (**Table 2**) and a PPV of 95% when it was tested against the gold standard definition in our second manual review method (**Table 3**). Despite this, PCOS_regex-strict_ algorithm had a sensitivity of only 50% and specificity of 29% when evaluated using the gold standard definition (**Table 3**).

When compared to our broader algorithms, the performance of the PCOS_regex-strict_ algorithm was most similar to the PCOS_coded-strict_ algorithm which had a slightly higher PPV of 98% (**Table 2**). The text mining criteria did not significantly improve the accuracy of PCOS_regex-strict_ over PCOS_coded-strict_. The accuracy of the “broad” criteria decreased compared to the “strict” algorithms. Our broadest algorithm, PCOS_coded-broad_, had the lowest performance with a PPV of 30% (**Table 2**). This is likely due to the large number of patients who only had an irregular menstruation ICD code with no other diagnosed PCOS symptoms. Interestingly, the addition of the text-mined PCOS keyword significantly improved the PPV of the broad criteria algorithm from 30% (PCOS_coded-broad_) to 82% (PCOS_regex-broad_). Unlike the extreme broad criteria, the other algorithms had a PCOS prevalence that reflected the prevalence observed in the population (18,27,28). This was also true for the race stratified cohorts where there were little to no differences in prevalence.

### Clinical feature characterization

We characterized the clinical profiles of cases and controls identified by each algorithm by calculating median BMI, insulin, estradiol, and free testosterone levels (**Table 4**). Each algorithm identified set was significantly heavier compared to controls (p < 0.05). For all algorithms, African American and Hispanic women had higher baseline metabolic profiles for BMI, insulin, estradiol, and free testosterone measurements compared to White women (1,2). Moreover, the median BMI in the algorithm datasets increased with increasing algorithm stringency (**Figure 2a**). African American women meeting algorithm criteria consistently demonstrated the highest median BMI in the PCOS_coded-broad_, PCOS_coded-strict_, and PCOS_regex-broad_ algorithms (**Supplementary Table 2**). The BMI of all three stratified race groups differed significantly from each other with the exception of White and Hispanic women in the strictest regex algorithms (**Figure 3a)**. Within the Hispanic defined group, the median values did not vary significantly between the PCOS_coded-strict_, PCOS_regex-broad_, and PCOS_regex-strict_ groups (**Supplementary Figure 3a**).

**Figure 2.**
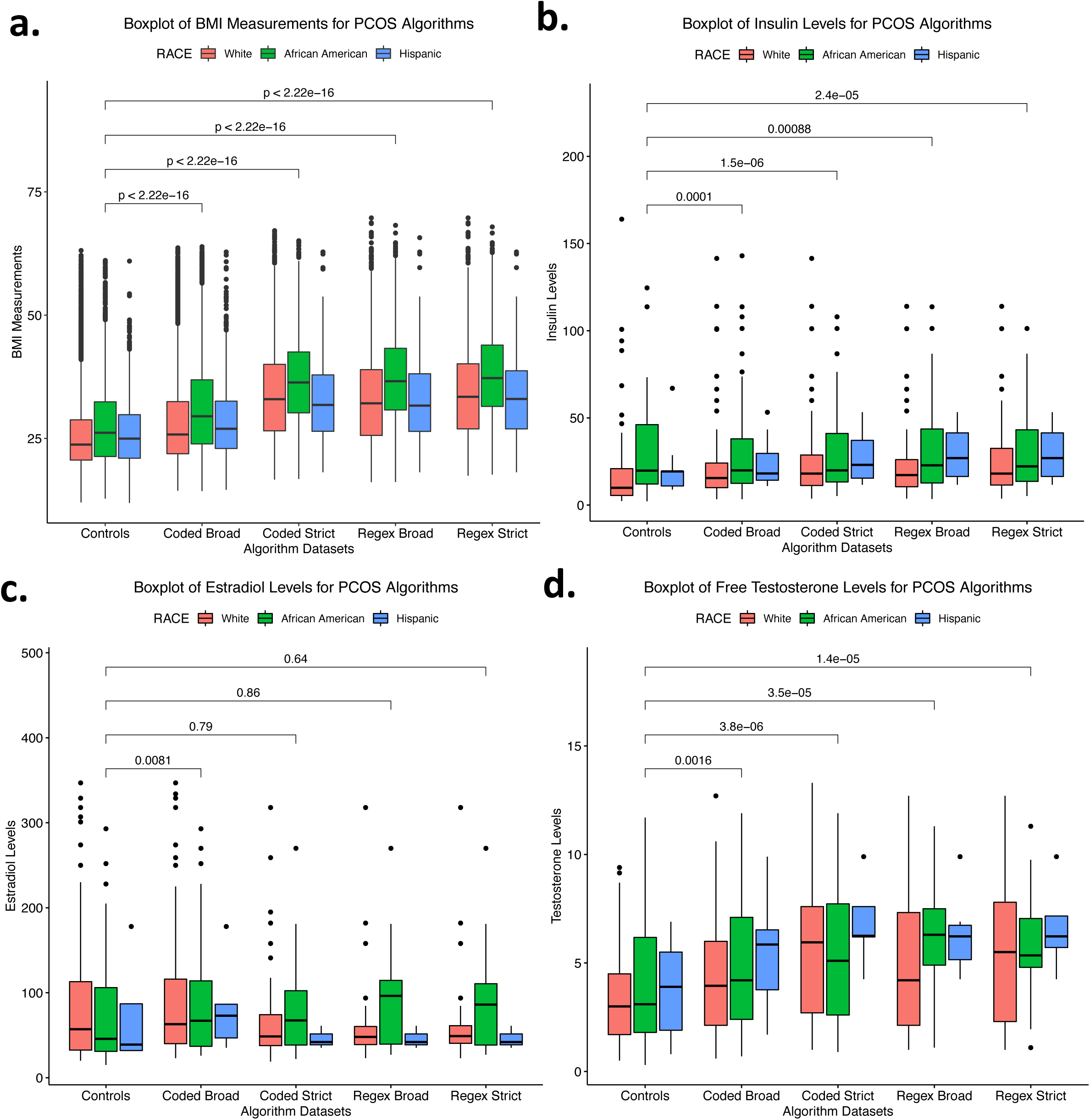
Laboratory measurements for PCOS algorithm cases and controls. Boxplots of (a) BMI measurements, (b) insulin levels, (c) estradiol levels, and (d) free testosterone levels for PCOS algorithm cases and controls. Colored boxes represent races. Boxes represent the individuals with lab measurements in the 25th and 75th percentile. Lines above and below the boxes represent the 95th and 5th quartiles. Lines within each box mark the median. Wilcoxon rank-sum tests were performed between algorithm cases and controls and Brackets display the p-values of the statistical test. BMI= body mass index.

**Figure 3.**
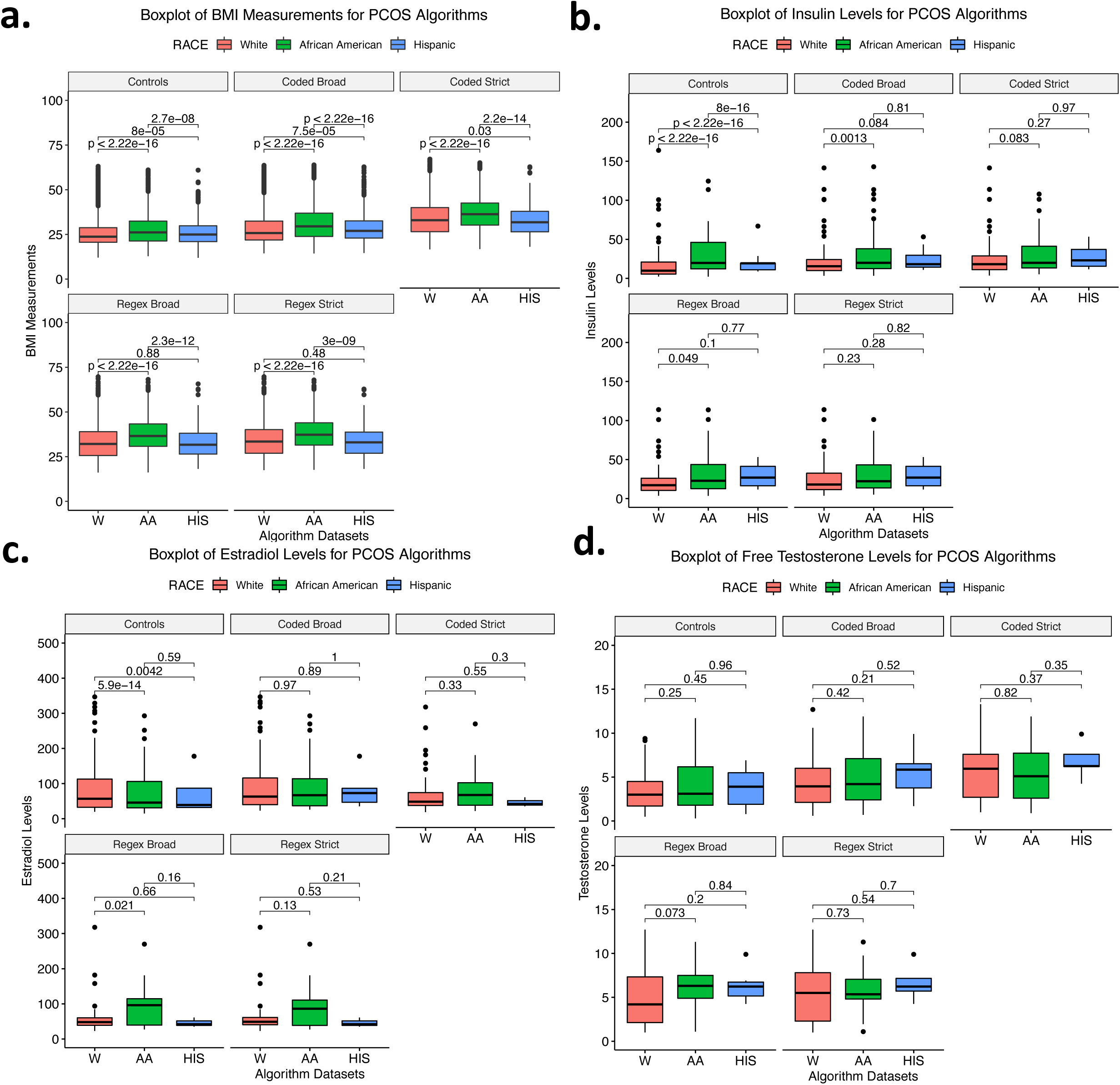
Comparison of laboratory measurements between White, African American, and Hispanic PCOS algorithm cases and controls. Boxplots of (a) BMI measurements, (b) insulin levels, (c) estradiol levels, and (d) free testosterone levels for White, African American, and Hispanic PCOS cases and controls. Colors correspond with each race. Boxes represent the 25th and 75th quartiles. Lines above and below the boxes represent the 95th and 5th quartiles. Lines within each box mark the median. Wilcoxon rank-sum tests were performed between races within the PCOS cases and control datasets. Brackets display the p-values of the statistical test. BMI= body mass index; W = White; AA = African American; HIS = Hispanic.

Overall, women identified by each of the four PCOS algorithms demonstrated significantly higher insulin and free testosterone levels compared to controls (p < 0.05) (**Figures 2b, 2d, Table 4**). The median hormone values also increased with algorithm stringency, though the trend was not as pronounced as it was for the BMI measurements. BMI increased with algorithm stringency within each race stratified analysis. Insulin values increased with algorithm stringency for White and Hispanic women, and free testosterone values increased with algorithm stringency for White and African American women (**Figure 4**, **Supplementary Table 2**).

**Figure 4.**
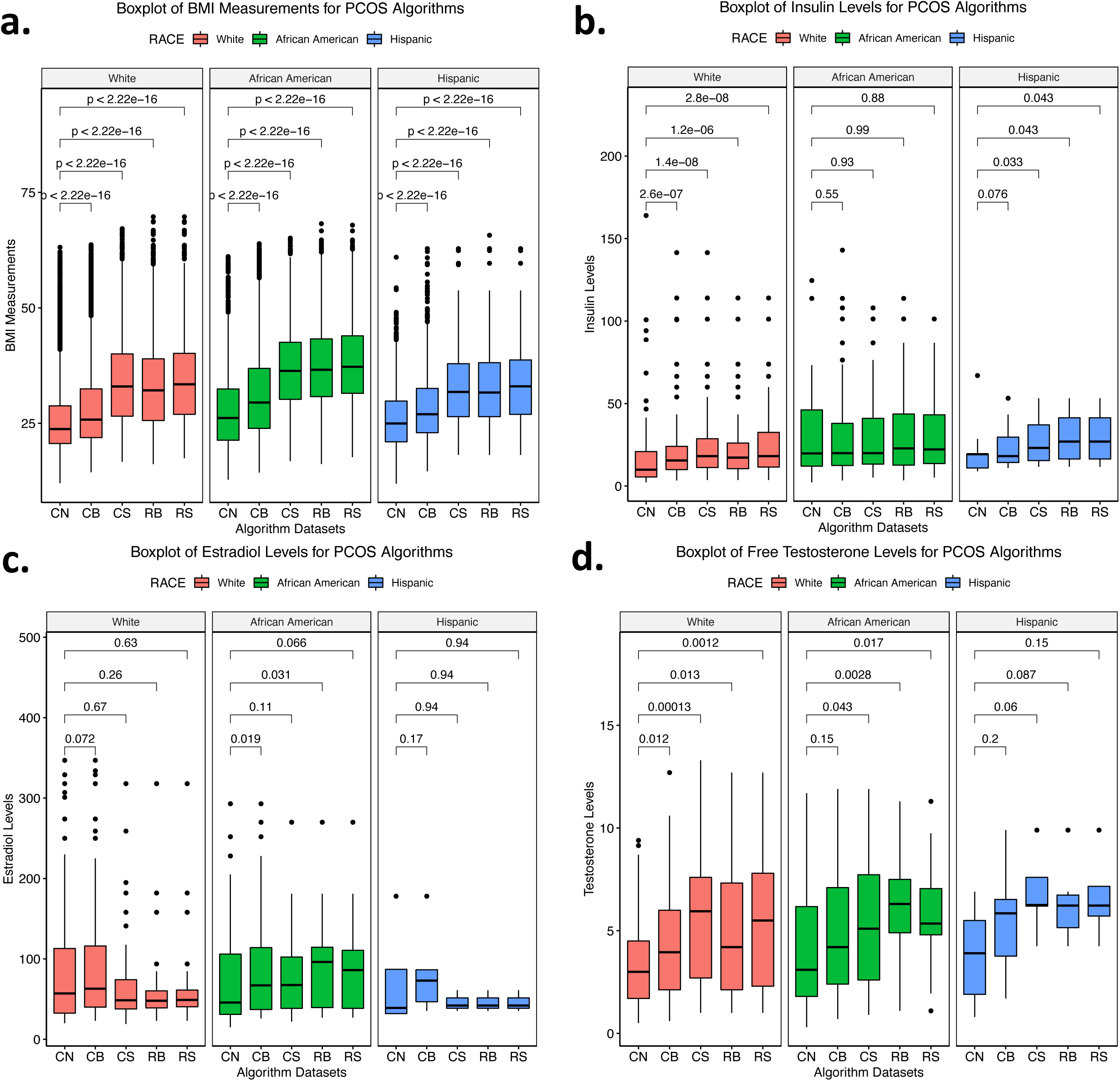
Race stratified laboratory measurements for PCOS algorithm cases and controls. Boxplots of (a) BMI measurements, (b) insulin levels, (c) estradiol levels, and (d) free testosterone levels of race stratified PCOS algorithm datasets. Each color corresponds with a race. Boxes represent the 25th and 75th quartiles. Lines above and below the boxes represent the 95th and 5th quartiles. Lines within each box mark the median. Wilcoxon rank-sum tests were performed between algorithm cases and controls within each race. Brackets display the p-values of the statistical test. BMI= body mass index; CN = controls; CB = coded broad; CS = coded strict; RB = regex broad; RS = regex strict.

Measurements of free testosterone were significantly higher among women identified by the PCOS_coded-strict_, PCOS_regex-broad_ and PCOS_regex-strict_ algorithms compared to the controls and the PCOS_coded-broad_ algorithm (**Supplementary Figure 2d**). The insulin profile of African Americans was similar across women identified by each algorithm and also did not significantly differ from controls (**Figure 4b**). However, African American women identified by the PCOS_coded-broad_ and PCOS_regex-broad_ algorithms did exhibit higher estradiol levels than African American controls (**Figure 4c**). Additionally, African American women identified by the PCOS_regex-broad_ algorithm demonstrated significantly higher estradiol levels than White women identified by the same approach (**Figure 3c**). Outside of these findings, there were no other significant differences between races for hormone values (**Figure 3**).

### Genetic validation of PCOS case status

Increasing algorithm requirement stringency was also reflected in the performance of the PCOS_PRS_. Odds ratios (ORs), which are reported per standard deviation of the PCOS_PRS_ were higher among cases identified by algorithms using strict PCOS requirements and decreased as those requirements were relaxed. This trend was observed for both the European and multi-ancestry genotyped samples (**Figure 5, Table 5**). BMI was tested as a covariate in the model and did not impact the results. (**Supplementary Table 3**).

**Figure 5.**
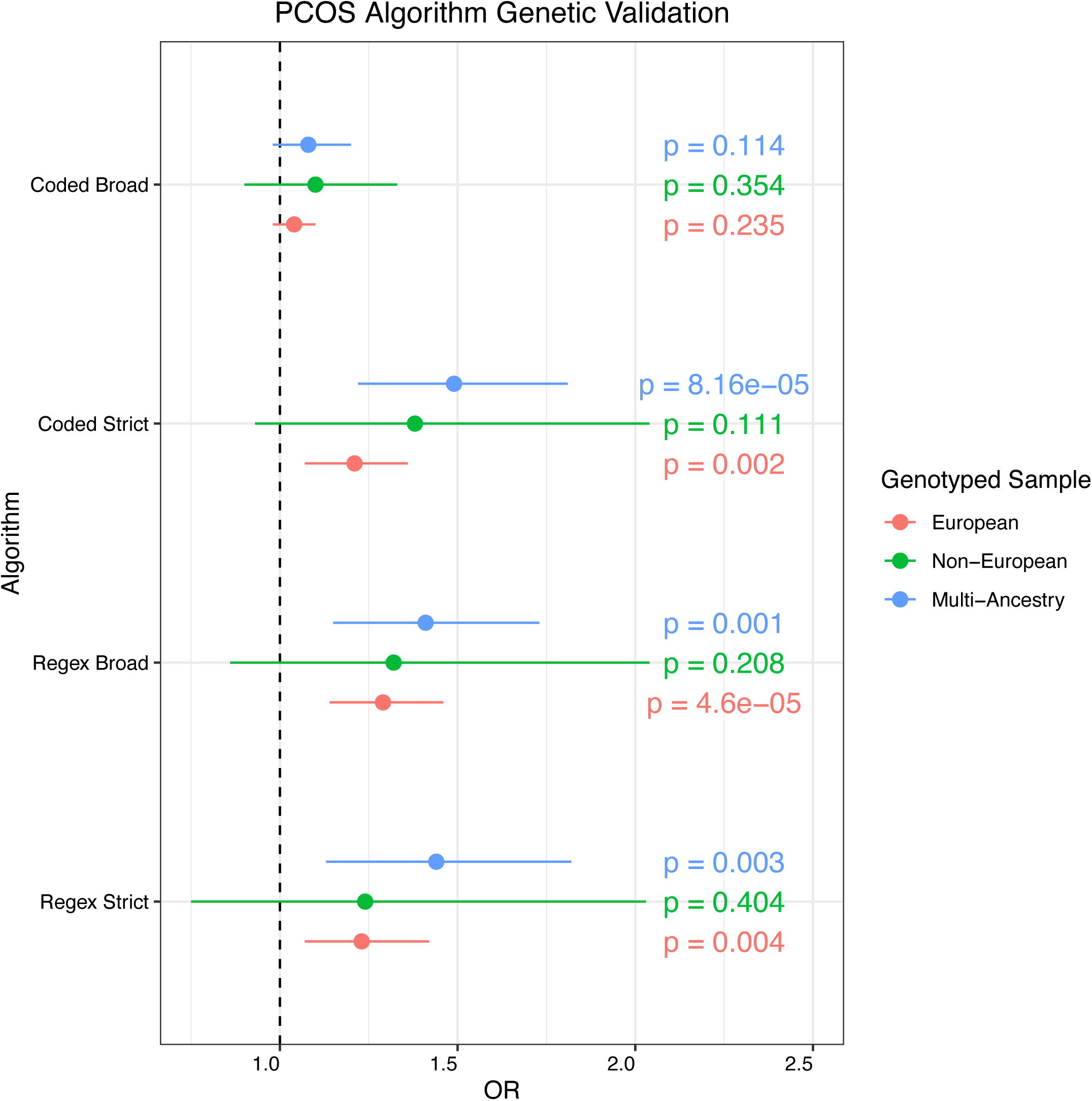
Genetic validation of PCOS algorithms. Logistic regressions were performed between PCOS case status and PCOS polygenic risk scores (PCOS_PRS_). The regression models were adjusted for median age and the first 10 principal components. P-values are displayed for each regression model. Colors correspond to the genotyped sample used in the analysis. OR = odds ratio.

Among European genotyped cases identified by the PCOS_regex-broad_ algorithm we observed an OR of 1.29 (p = 4.6E-5) and the greatest variance explained in case/control status by the PCOS_PRS_ model (pseudo-r^2^ = 0.83%, p = 4.6E-5) (**Table 5**). When the PCOS_PRS_ model was applied to the multi-ancestry sample, the PCOS_coded-broad_ yielded the highest OR (OR = 1.49, p = 8.7E-5), followed by PCOS_regex-strict_ (OR = 1.44, p = 0.003) and PCOS_regex-broad_ (OR = 1.41, p = 0.001). The PCOS_coded-strict_ algorithm (pseudo-r^2^ = 0.7%, p = 8.7E-5) also had the most variance explained by the model.

Similar to the European genotyped sample, only the PCOS_regex-broad_ and the PCOS_coded-strict_ algorithm passed at least one of our two multiple correction tests.

There were no significant associations between PCOS_PRS_ and case/control status within the non-European genotyped sample. We are underpowered in this analysis, which is evidenced by large confidence intervals in comparison to the European and multi-ancestry genotyped sample (**Figure 5, Table 5**).

### Association between PCOS_PRS_ and hormone measurements

There was no association between PCOS_PRS_ and any of the hormone measures (insulin, estradiol, and free testosterone) evaluated. (**Supplementary Tables 4-6**).

## Discussion

Expanding study inclusion criteria has the potential to increase and diversify future study samples which may significantly benefit underrepresented high risk groups (27). This study describes an automated PCOS phenotyping method that captures varying degrees of PCOS symptoms, parallel dysregulated hormone levels, and increased genetic risk for the PCOS_coded-strict_, PCOS_regex-broad_, and PCOS_regex-strict_ algorithms. Our results also further support the hypothesis that women with PCOS symptoms have increased metabolic dysregulation compared to controls when measured on BMI, insulin, and free testosterone values dependent on clinical definitions and their EHR-reported race. Finally, our study demonstrates the importance of including multiethnic genotyped samples in addition to European populations, for genetic analysis.

Our two “strict definition” algorithms identified PCOS cases with high accuracy (PPV>95%) and the PCOS_regex-strict_ algorithm performed just as well when compared to the gold standard dataset (PPV = 95%). Although the sensitivity and the specificity of the PCOS_regex-strict_ algorithm was low, the high PPV displays its high accuracy within an EHR setting. Compared to the previously reported ICD based PCOS algorithm, which had a PPV of 66% and 65% for their “broad” and “refined” algorithm, our two best performing algorithms improved case identification by over 30% (9). Both the Castro et al. study and our study show the feasibility of using ICD based algorithms to identify PCOS cases and the advantages of using EHR systems. Consistent with the previous study, we also found that inclusion of text mining increased the proportion of true PCOS cases (PPV = 98%) but resulted in a smaller sample size. Removal of the text mining requirement only slightly impacted the PPV (96%), indicating that studies performed in EHR systems with limited access to clinical notes and ultrasound images may still be able to identify women with PCOS with high accuracy. Conversely, we found that the addition of text mining to the broad algorithms significantly improved the accuracy from 30% to 82%, indicating that clinical suspicions of PCOS documented in medical records may also be used to identify women with PCOS even when few symptoms are coded.

The clinical and epidemiological features of our PCOS cohorts are consistent with previous studies that show women with PCOS have higher metabolic clinical characteristics than controls (29-31). We also observed no significant difference in the prevalence of PCOS definitions between minority and White individuals, nor were there any large differences between races for insulin, estradiol, and free testosterone clinical measurements (32). However, African American had significantly higher BMI measurements than Hispanic and White women across both the control and PCOS definition datasets. African Americans are disproportionately affected by conditions such as obesity and type 2 diabetes, which could possibly be exacerbated by the onset of PCOS (33,34). These conditions could also be influenced by the patient structure of the EHR database which will have a sicker population due to the tertiary care status of VUMC and due to its geographic location where obesity rates are generally higher.

Polygenic risk scores provide the unique ability to examine the genetic susceptibility of PCOS across different clinical definitions and ancestries. However, it is important to note that, as of to date, there are no PCOS GWAS in African descent populations, and that PRS built from Eurocentric populations will not perform as robustly in non-European samples (35). Despite this, previous studies have shown that PCOS_PRS_ models can detect cross-ancestry genetic risk in African and multi-ancestry cohorts using European based PRS models (36). In our study, the PCOS_PRS_ model explained little variance in case/control status (pseudo-r^2^ < 1%), but the PCOS_PRS_ was significantly higher among cases in the PCOS_coded-strict_, PCOS_regex-broad_, and the PCOS_regex-strict_ algorithms compared to controls.

Eurocentric GWAS remain a significant limitation to interpretability of PRS in non-European samples and studies including genetic data gathered on diverse ancestral populations are needed to advance PCOS genomics in the EHR (35,37). The limitations of our study include low statistical power in the genetic analysis of non-European cases and controls. This limitation is primarily a function of the under-representation of non-European populations in the PCOS GWAS, but also reflects the low representation of non-European samples in the VUMC EHR.

In conclusion, we applied an automated phenotyping approach to examine variation in the symptomology of PCOS across racial and ethnic groups using both strict and broad classification criteria. Debate over current implemented PCOS diagnostic criteria, in part, reflects the complexity of PCOS symptoms and EHRs provide significant advantages. First, research in the longitudinal EHR allows us to maximize the number of PCOS clinical manifestations observed over time (16,28,38). Secondly, EHRs contain a variety of clinical variables (e.g., laboratory measurements, imaging records, etc.) that can be used to study the risk factors and health outcomes of PCOS patients. We created a simple EHR-based PCOS algorithm that identifies patients using ICD codes and text mining (PCOS_regex-strict_) methods and by relaxing the inclusion requirements we identified patients across the PCOS syndromic spectrum. This study approach provides a unique view of the shared clinical and genetic architecture that influences both PCOS as a discrete diagnosis, and the symptoms of PCOS that affect a large number of women.

## Data Availability

Due to data sharing restrictions related to privacy concerns in the EHR, the datasets generated from our hospital population will not be publicly available, however, all criteria for automated phenotyping is available in supplementary materials.

## Funding/Support

KVA and JC are supported by NIH training grant 5T32GM007628-42 and 5T32DK007061. LKD is supported by U54MD010722.This research was done in part using the resources from the Advanced Computing Center for Research and Education at Vanderbilt University, Nashville, TN. Datasets for this project were obtained using the Synthetic Derivative at Vanderbilt University Medical Center which is supported by multiple grant sources that are institutional, private, and federal. This includes the NIH funded Shared Instrumentation Grant S10RR025141 and CTSA grants UL1TR002243, UL1TR000445, and UL1RR024975.

## Authors contributions

KVA and LKD drafted the manuscript. KVA, DH, and KS performed chart review. JC and MW supervised and reviewed chart review. All authors contributed to the interpretation of the data and the revision of the manuscript.

## Disclosures

The authors have nothing to disclose.

## Notes

### Competing Interest Statement

The authors have declared no competing interest.

## References

1. Wolf WM, Wattick RA, Kinkade ON, Olfert MD. Geographical prevalence of polycystic ovary syndrome as determined by region and race/ethnicity. Int. J. Environ. Res. Public Health 2018;15(11):2589.

2. Teede H, Gibson-Helm M, Norman RJ, Boyle J. Polycystic Ovary Syndrome: Perceptions and Attitudes of Women and Primary Health Care Physicians on Features of PCOS and Renaming the Syndrome. J Clin Endocrinol Metab 2014;(1):99.

3. Eshre TR, Group A-SPCW. Revised 2003 consensus on diagnostic criteria and long-term health risks related to polycystic ovary syndrome. Fertil. Steril. 2004;81(1):19–25.

4. Azziz R, Carmina E, Dewailly D, Diamanti-Kandarakis E, Escobar-Morreale F, Futterweit W, Janssen OE, Legro RS, Norman RJ, Taylor AE, Witchel SF, (Task Force on the Phenotype of the Polycystic Ovary Syndrome of The Androgen Excess and PCOS Society)*, Ector H, Escobar-Morreale F, Futterweit W, Janssen OE, Legro RS, Norman RJ, Taylor AE, Witchel SF. The Androgen Excess and PCOS Society criteria for the polycystic ovary syndrome: the complete task force report. Fertil. Steril. 2009;91(2):456–488.

5. Indran IR, Huang Z, Khin LW, Chan JK, Viardot-Foucault V, Yong EL. Simplified 4-item criteria for polycystic ovary syndrome: A bridge too far? Clin. Endocrinol. (Oxf). 2018;89(2):202–211.

6. Stepto NK, Cassar S, Joham AE, Hutchison SK, Harrison CL, Goldstein RF, Teede HJ. Women with polycystic ovary syndrome have intrinsic insulin resistance on euglycaemic-hyperinsulaemic clamp. Hum. Reprod. 2013;28(3):777–784.

7. Hickey M, Doherty DA, Atkinson H, Sloboda DM, Franks S, Norman RJ, Hart R. Clinical, ultrasound and biochemical features of polycystic ovary syndrome in adolescents: Implications for diagnosis. Hum. Reprod. 2011;26(6):1469–1477.

8. Copp T, Jansen J, Doust J, Mol BW, Dokras A, McCaffery K. Are expanding disease definitions unnecessarily labelling women with polycystic ovary syndrome? BMJ 2017;358. doi:10.1136/bmj.j3694.

9. Castro V, Shen Y, Yu S, Finan S, Pau CT, Gainer V, Keefe CC, Savova G, Murphy SN, Cai T, Welt CK. Identification of subjects with polycystic ovary syndrome using electronic health records. Reprod. Biol. Endocrinol. 2015;13(1):1–8.

10. Jacewicz-Święcka M, Kowalska I. Polycystic ovary syndrome and the risk of cardiometabolic complications in longitudinal studies. Diabetes. Metab. Res. Rev. 2018;34(8). doi:10.1002/dmrr.3054.

11. Pavicic Baldani D, Skrgatic L, Ougouag R. Polycystic Ovary Syndrome: Important Underrecognised Cardiometabolic Risk Factor in Reproductive-Age Women. 2015. doi:10.1155/2015/786362.

12. Chan JL, Kar S, Vanky E, Morin-Papunen L, Piltonen T, Puurunen J, Tapanainen JS, Maciel GAR, Hayashida SAY, Soares Jr JM. Racial and ethnic differences in the prevalence of metabolic syndrome and its components of metabolic syndrome in women with polycystic ovary syndrome: a regional crosssectional study. Am. J. Obstet. Gynecol. 2017;217(2):189. e1–189. e8.

13. Engmann L, Jin S, Sun F, Legro RS, Polotsky AJ, Hansen KR, Coutifaris C, Diamond MP, Eisenberg E, Zhang H, Santoro N. Racial and ethnic differences in the polycystic ovary syndrome metabolic phenotype. 2017. doi:10.1016/j.ajog.2017.01.003.

14. Meyer ML, Sotres-Alvarez D, Steiner AZ, Cousins L, Talavera GA, Cai J, Daviglus ML, Loehr LR. Polycystic ovary syndrome signs and metabolic syndrome in premenopausal Hispanic/Latina women: The HCHS/SOL study. J. Clin. Endocrinol. Metab. 2020. doi:10.1210/clinem/dgaa012.

15. Hillman JK, Johnson LNC, Limaye M, Feldman RA, Sammel M, Dokras A. Black women with polycystic ovary syndrome (PCOS) have increased risk for metabolic syndrome and cardiovascular disease compared with white women with PCOS. Fertil. Steril. 2014;101(2):530–535.

16. March WA, Moore VM, Willson KJ, Phillips DIW, Norman RJ, Davies MJ. The prevalence of polycystic ovary syndrome in a community sample assessed under contrasting diagnostic criteria. Hum. Reprod. 2010;25(2):544–551.

17. Ding T, Hardiman PJ, Petersen I, Wang F-F, Qu F, Baio G. The prevalence of polycystic ovary syndrome in reproductive-aged women of different ethnicity: a systematic review and meta-analysis. Oncotarget 2017;8(56):96351–96358.

18. Bozdag G, Mumusoglu S, Zengin D, Karabulut E, Yildiz BO. The prevalence and phenotypic features of polycystic ovary syndrome: A systematic review and meta-Analysis. Hum. Reprod. 2016;31(12):2841–2855.

19. Robinson JR, Wei W-Q, Roden DM, Denny JC. Defining phenotypes from clinical data to drive genomic research. Annu. Rev. Biomed. Data Sci. 2018;1:69–92.

20. Dennis JK, Sealock JM, Straub P, Hucks D, Actkins K, Faucon A, Goleva SB, Nirachou M, Singh K, Morley T, Ruderfer DM, Mosley JD, Chen G, Davis LK. Lab-wide association scan of polygenic scores identifies biomarkers of complex disease. *medRxiv* 2020:2020.01.24.20018713.

21. Bien SA, Wojcik GL, Zubair N, Gignoux CR, Martin AR, Kocarnik JM, Martin LW, Buyske S, Haessler J, Walker RW, Cheng I, Graff M, Xia L, Franceschini N, Matise T, James R, Hindorff L, Le Marchand L, North KE, Haiman CA, Peters U, Loos RJF, Kooperberg CL, Bustamante CD, Kenny EE, Carlson CS. Strategies for Enriching Variant Coverage in Candidate Disease Loci on a Multiethnic Genotyping Array. PLoS One 2016;11(12):167758.

22. Price AL, Patterson NJ, Plenge RM, Weinblatt ME, Shadick NA, Reich D. Principal components analysis corrects for stratification in genome-wide association studies. Nat. Genet. 2006;38(8):904–909.

23. Chang CC, Chow CC, Tellier LC, Vattikuti S, Purcell SM, Lee JJ. Second-generation PLINK: rising to the challenge of larger and richer datasets. Gigascience 2015;4(7):7.

24. Das S, Forer L, Schönherr S, Sidore C, Locke AE, Kwong A, Vrieze SI, Chew EY, Levy S, McGue M, Schlessinger D, Stambolian D, Loh PR, lacono WG, Swaroop A, Scott LJ, Cucca F, Kronenberg F, Boehnke M, Abecasis GR, Fuchsberger C. Next-generation genotype imputation service and methods. Nat. Genet. 2016;48(10):1284–1287.

25. Ge T, Chen C-Y, Ni Y, Feng Y-CA, Smoller JW. Polygenic prediction via Bayesian regression and continuous shrinkage priors. Nat. Commun. 2019;10(1776):1–10.

26. Day F, Karaderi T, Jones MR, Meun C, He C, Drong A, Kraft P, Lin N, Huang H, Broer L. Large-scale genome-wide meta-analysis of polycystic ovary syndrome suggests shared genetic architecture for different diagnosis criteria. PLoS Genet. 2018;14(12):e1007813.

27. Neven ACH, Laven J, Teede HJ, Boyle JA. A summary on polycystic ovary syndrome: Diagnostic criteria, prevalence, clinical manifestations, and management according to the latest international guidelines. Semin. Reprod. Med. 2018;36(1):5–12.

28. Skiba MA, Islam RM, Bell R, Davis SR. Understanding variation in prevalence estimates of polycystic ovary syndrome: a systematic review and meta-analysis. Hum. Reprod. Update 2018;7(1). doi:10.1093/humupd/7.1.1.

29. Lerchbaum E, Schwetz V, Rabe T, Giuliani A, Obermayer-Pietsch B. Hyperandrogenemia in Polycystic Ovary Syndrome: Exploration of the Role of Free Testosterone and Androstenedione in Metabolic Phenotype. 2014. doi:10.1371/journal.pone.0108263.

30. Antonio L, Pauwels S, Laurent MR, Vanschoubroeck D, Jans I, Billen J, Claessens F, Decallonne B, De Neubourg D, Vermeersch P, Vanderschueren D. Free Testosterone Reflects Metabolic as well as Ovarian Disturbances in Subfertile Oligomenorrheic Women. Int. J. Endocrinol. 2018. doi:10.1155/2018/7956951.

31. Diamanti-Kandarakis E, Dunaif A. Insulin Resistance and the Polycystic Ovary Syndrome Revisited: An Update on Mechanisms and Implications. 2012. doi:10.1210/er.2011-1034.

32. Ladson G, Dodson WC, Sweet SD, Archibong AE, Kunselman AR, Demers LM, Williams NI, Coney P, Legro RS. Racial Influence on the Polycystic Ovary Syndrome Phenotype: A Black White Case-Control Study. Fertil Steril 2011;96(1):224–229.

33. Huddleston HG, Cedars MI, Sohn SH, Giudice LC, Fujimoto VY. Racial and ethnic disparities in reproductive endocrinology and infertility. Am. J. Obstet. Gynecol. 2010;202(5):413–419.

34. Butts SF, Seifer DB. Racial and ethnic differences in reproductive potential across the life cycle. Fertil. Steril. 2010;93(3):681–690.

35. Martin AR, Kanai M, Kamatani Y, Okada Y, Neale BM, Daly MJ. Clinical use of current polygenic risk scores may exacerbate health disparities. 2019;51(4):584–591.

36. Joo YY, Actkins K, Pacheco JA, Basile AO, Carroll R, Crosslin DR, Day F, Denny JC, Velez Edwards DR, Hakonarson H, Harley JB, Hebbring SJ, Ho K, Jarvik GP, Jones M, Karderi T, Mentch FD, Meun C, Namjou B, Pendergrass S, Ritchie MD, Stanaway IB, Urbanek M, Walunas TL, Smith M, Chisholm RL, Kho AN, Davis L, Hayes MG. A polygenic and phenotypic risk prediction for Polycystic Ovary Syndrome evaluated by Phenome-wide association studies. J. Clin. Endocrinol. Metab. 2020. doi:10.1210/clinem/dgz326.

37. Wojcik GL, Graff M, Nishimura KK, Tao R, Haessler J, Gignoux CR, Highland HM, Patel YM, Sorokin EP, Avery CL, Belbin GM, Bien SA, Cheng I, Cullina S, Hodonsky CJ, Hu Y, Huckins LM, Jeff J, Justice AE, Kocarnik JM, Lim U, Lin BM, Lu Y, Nelson SC, Park S-SLSL, Poisner H, Preuss MH, Richard MA, Schurmann C, Setiawan VW, Sockell A, Vahi K, Verbanck M, Vishnu A, Walker RW, Young KL, Zubair N, Acuña-Alonso V, Ambite JL, Barnes KC, Boerwinkle E, Bottinger EP, Bustamante CD, Caberto C, Canizales-Quinteros S, Conomos MP, Deelman E, Do R, Doheny K, Fernández-Rhodes L, Fornage M, Hailu B, Heiss G, Henn BM, Hindorff LA, Jackson RD, Laurie CACC, Laurie CACC, Li Y, Lin D-YY, Moreno-Estrada A, Nadkarni G, Norman PJ, Pooler LC, Reiner AP, Romm J, Sabatti C, Sandoval K, Sheng X, Stahl EA, Stram DO, Thornton TA, Wassel CL, Wilkens LR, Winkler CA, Yoneyama S, Buyske S, Haiman CA, Kooperberg C, Le Marchand L, Loos RJFF, Matise TC, North KE, Peters U, Kenny EE, Carlson CS. Genetic analyses of diverse populations improves discovery for complex traits. Nature Publishing Group; 2019: 514–518.

38. Wang S, Alvero R. Racial and ethnic differences in physiology and clinical symptoms of polycystic ovary syndrome. Semin. Reprod. Med. 2013;31(5):365–369.

